# *GBA1* variants with unknown classification are modest contributors to Parkinson’s disease susceptibility

**DOI:** 10.1101/2025.09.05.25335124

**Authors:** Sitki Cem Parlar, Yoomin Lee, Ziv Gan-Or

**Affiliations:** Department of Human Genetics, McGill University, Montréal, Québec, Canada; The Neuro (Montreal Neurological Institute-Hospital), McGill University, Montréal, Québec, Canada; Department of Neurology and Neurosurgery, McGill University, Montréal, Québec, Canada

## Abstract

**Background:** *GBA1* variants cause Gaucher disease (GD) in biallelic forms and increase Parkinson’s disease (PD) risk in heterozygous carriers. Carriers of ‘severe’ or ‘mild’ variants (causing GD type 1 or types 2-3) can enroll in clinical trials, whereas those with ‘unknown’ variants are typically excluded.

**Objectives:** We assessed the contribution of ‘unknown’ variants to PD risk and their relevance for trial stratification.

**Methods:** We meta-analyzed 34 case–control studies (24,060 PD cases, 14,465 controls). Odds ratios (OR) were estimated using random-effects models and stratified by the American College of Medical Genetics (ACMG) criteria.

**Results:** ‘Unknown’ variants also classified as variants of uncertain significance (VUS) per ACMG criteria were associated with PD (OR=1.59, 95%CI:1.25–2.02; I^2^=0%). VUS + likely pathogenic + pathogenic also showed association (OR=1.63, 95%CI:1.28–2.06; I^2^=0%).

**Conclusions:** ‘Unknown’ *GBA1* variants may be considered in clinical trials if also classified as VUS, likely pathogenic, or pathogenic per ACMG criteria.

## INTRODUCTION

*GBA1* variants are amongst the most common genetic risk factors of Parkinson’s disease (PD), with a prevalence of 5-20% in PD patients of different ethnicities. Numerous clinical trials in various stages are thus targeting glucocerebrosidase, the enzyme encoded by *GBA1*. One critical aspect of these trials’ design is the classification of *GBA1* variants, which can affect progression rate of PD and therefore trial outcomes.^1^

Currently, *GBA1* variants are classified based on their association with PD and Gaucher disease (GD), a lysosomal storage disorder caused by biallelic *GBA1* variants. GD can be classified into three types: 1) GD type 1, a mild form of GD with no central nervous system involvement 2) GD type 2, a severe form of GD with brain involvement and death in early childhood, and 3) GD type 3, an intermediate-severe form which also includes brain involvement, but patients can live into adulthood. *GBA1* variants can be classified accordingly: ‘mild’ *GBA1* variants are those that cause GD type 1, and ‘severe’ *GBA1* variants cause GD type 2 or 3. To be able to classify a *GBA1* variant, it therefore must be either reported in GD in a homozygous form or coupled with a known severe mutation. An additional class of variants is ‘risk’ variants, which are those that do not cause GD, yet are proven to be risk factors of PD. Currently, only two variants, p.E326K and p.T369M, can be reliably classified as ‘risk’ variants.^2^

This classification for ‘risk’, ‘mild’ and ‘severe’ variants has been shown to be clinically meaningful. Carriers of risk variants have an OR of below 2 to have PD,^3, 4^ while in carriers of ‘mild’ mutations it is typically an OR of 2-3, and ‘severe’ variants with an OR >5.^5^ The different classes of variants may also affect PD progression; ‘severe’ variant carriers seem to have faster motor and non-motor progression, and reduced survival.^6-8^ Therefore, when designing clinical trials targeting carriers of *GBA1* variants, it is important to balance the treatment and placebo arms of the trial according to this classification. If one arm of the trial has more ‘severe’ *GBA1* variant carriers than the other, that arm might progress faster than the other arm and affect trial outcomes.

However, numerous *GBA1* variants remain with an ‘unknown’ classification, as they have not been observed at a homozygous state or with a known severe variant in GD.^2^ Currently individuals with ‘unknown’ variants are typically excluded for *GBA1*-targeting trials. The goal of the current study is to quantify the contribution of this class of variants to PD risk.

## METHODS

### Data collection

We collected a list of all published studies reporting *GBA1* variants in PD cohorts, relying on the *GBA1*-PD browser.^2^ This open-access resource, which aggregates published studies and provides curated variant annotations, was accessed in its most recent version (October 2024). For the present analysis, we restricted inclusion to case–control studies reporting nonsynonymous coding *GBA1* variants with ‘unknown’ classification. We found 34 studies that reported *GBA1* variants with unknown classification, comprising 24,060 PD cases and 14,465 controls (see Supplementary Data 1).

### Variant nomenclature and classification

Variant annotations were obtained directly from the *GBA1*-PD browser.^2^ Nomenclature for *GBA1* variants was reported both with and without adjustment for the 39 amino acids cleaved at the N-terminus, to account for differences in reporting conventions. The browser also provides harmonized classifications based on two parallel systems: clinical GD severity and the American College of Medical Genetics and Genomics (ACMG) framework.^9^ The GD-based categories include ‘mild’ (variants causing GD type I), ‘severe’ (variants causing GD type II or III), ‘risk’ variants (variants not causing GD but associated with PD risk, namely p.E326K and p.T369M), and ‘unknown’. The ACMG framework provides a standardized system for evaluating the clinical significance of genetic variants, classifying them as pathogenic, likely pathogenic, variant of uncertain significance (VUS), likely benign, or benign.^9^ These two classification standards formed the basis of all downstream analyses.

### Meta-analysis

For each study, odds ratios (ORs) with 95% confidence intervals (CIs) were calculated from 2×2 contingency tables comparing the number of carriers of *GBA1* variants with unknown GD status among patients with PD and controls. The standard error of the log(OR) was derived using the inverse variance method. To account for variability across studies, pooled estimates were obtained using a random-effects model. Between-study heterogeneity was assessed using Cochran’s Q statistic and quantified by the I^2^ index. A constant continuity correction of 0.5 was applied to allow OR calculations where zero counts were present in the case or control arms of a study. Forest plots were generated in Python using the matplotlib package.^10^

## DATA SHARING

All information relevant to the analyses in this study, including the full list of variants examined and the studies considered, is available in the supplementary files of this article.

## RESULTS

We identified 125 unique nonsynonymous variants in the *GBA1* gene with ‘unknown’ classification. Based on the ACMG classification of these variants, 2 were pathogenic, 7 were likely pathogenic, 111 were variants of uncertain significance, and 5 were likely benign. The complete list of variants per study, including study-level case and control counts and ACMG classifications, is presented in Supplementary Data 2. Overall, we found 307 (1.28%) unknown variants in PD patients and 138 (0.95%) variants in controls.

We first analyzed all *GBA1* variants classified as ‘unknown’ per study and performed a random-effects meta-analysis (Figure 1). There was no statistically significant association in the meta-analysis (pooled OR = 1.34, 95% CI: 0.93–1.93, p = 0.12), and there was evidence of moderate heterogeneity (Q = 69.3, df = 33, p = 2.2×10□□; I^2^ = 52.4%). However, when variants were further stratified by ACMG classification, variants classified as VUS were associated with PD (OR=1.59, 95% CI: 1.25–2.02, p = 1.58 × 10^-4^, Figure 2A), with no evidence of heterogeneity (Q = 27.5, df = 30, p = 0.60; I^2^ = 0%). When VUS, likely pathogenic, and pathogenic variants were analyzed together (Figure 2B), the pooled OR was 1.63 (95% CI: 1.28– 2.06, p = 6.12× 10^-5^), also without evidence of heterogeneity (Q = 28.0, df = 30, p = 0.57; I^2^ = 0%). Analyses of likely pathogenic and pathogenic variants separately were not feasible due to insufficient sample size (data not shown).

**Figure 1.**
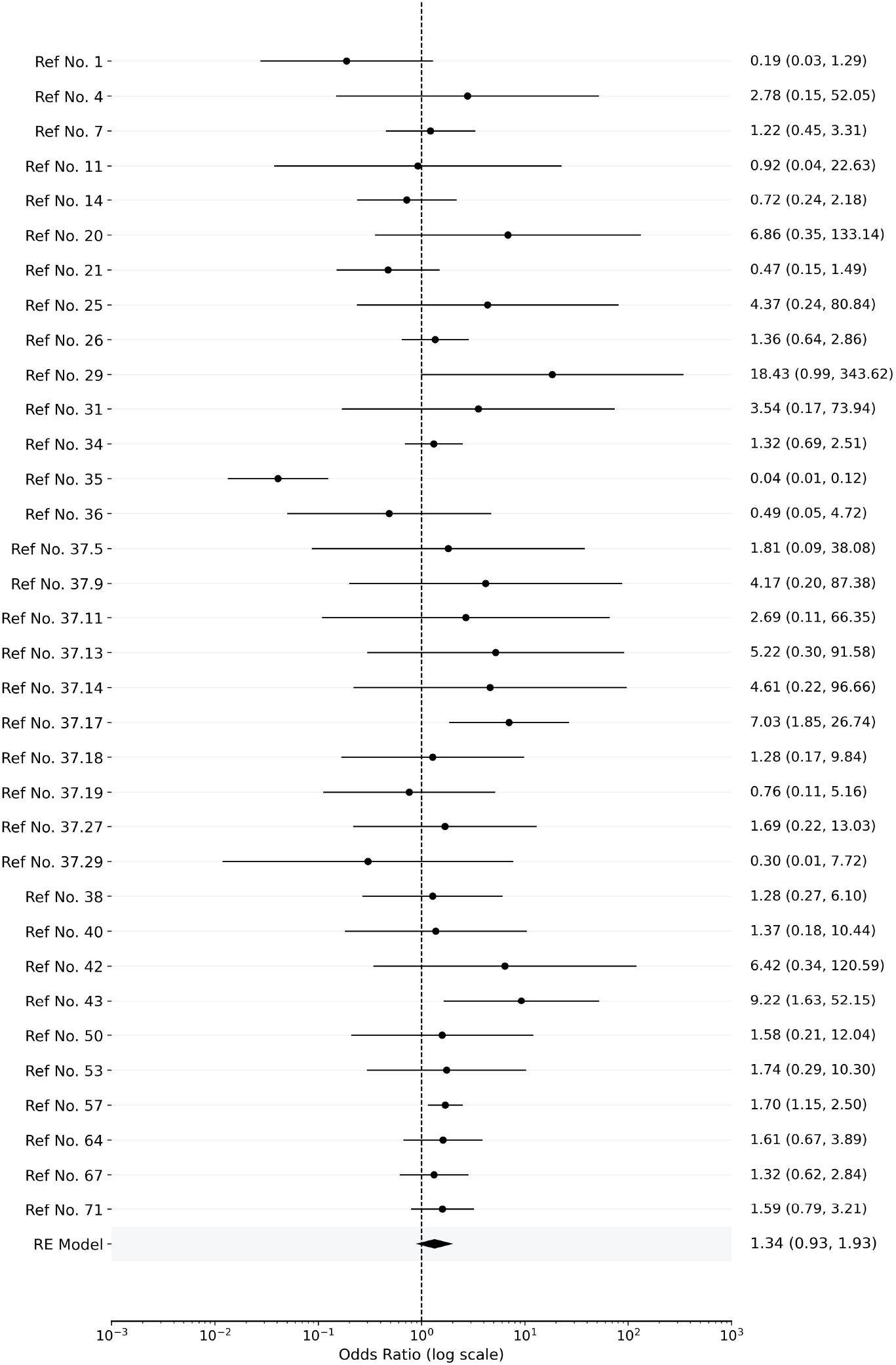
Forest plot of studies with data on *GBA1* variants classified as ‘unknown’. RE = random effects; Ref No. = *GBA1*-PD browser reference number.

**Figure 2.**
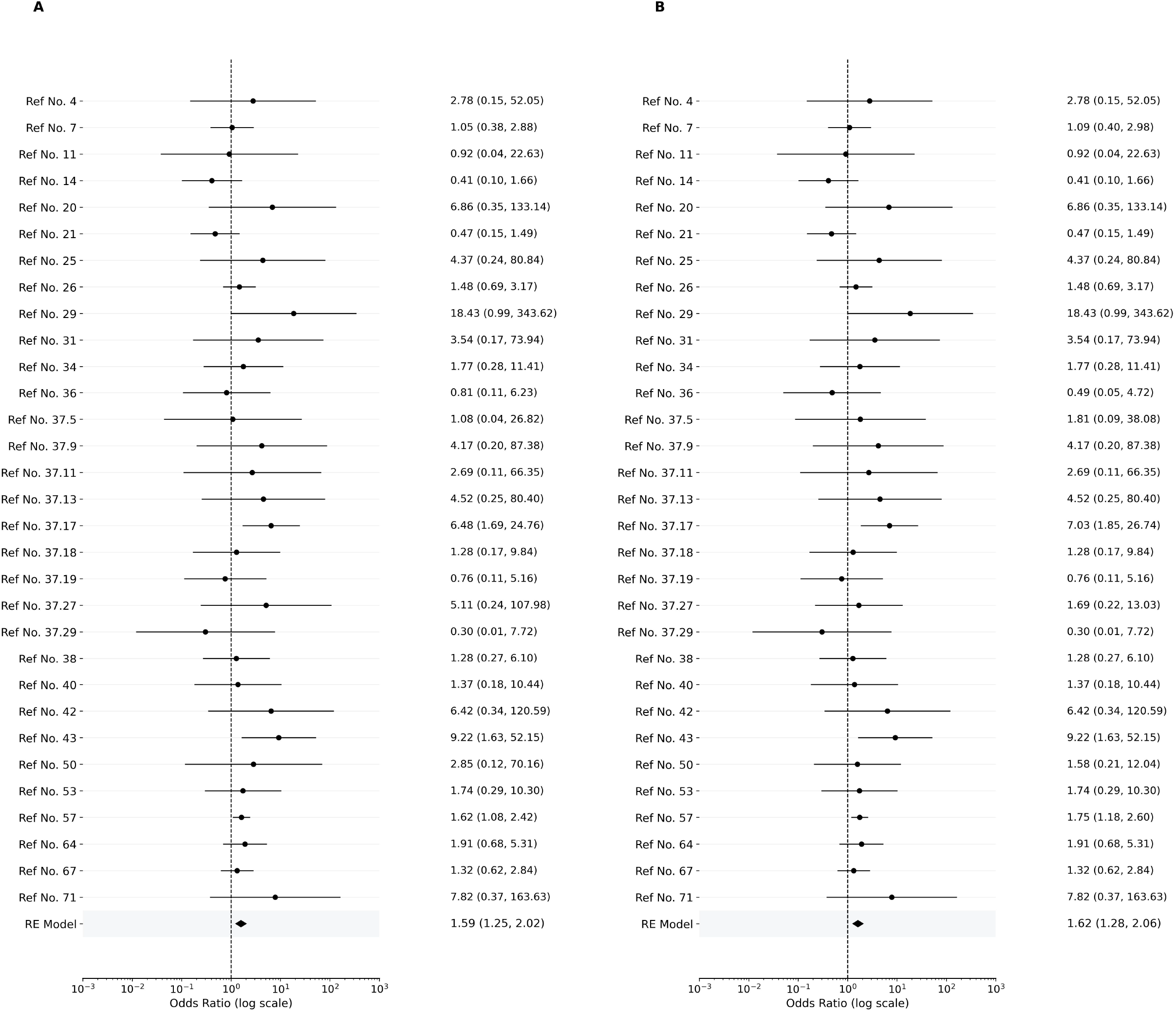
(A) Forest plot of studies with *GBA1* variants classified as ‘unknown’ and ACMG VUS. (B) Forest plot of studies with *GBA1* variants classified as ‘unknown’ and as ACMG VUS, likely pathogenic, or pathogenic.

Lastly, we ran single-variant OR calculations and meta-analysis for p.K(−27)R (pooled OR = 0.53, 95% CI: 0.12–2.35, p = 0.40; see Supplemental Figure 1) and p.E388K (pooled OR = 1.36, 95% CI: 0.48–3.89, p = 0.57; see Supplemental Figure 2).

## DISCUSSION

Our results show that *GBA1* variants classified as ‘unknown’ based on their contribution to GD, have a modest contribution to PD susceptibility. This is especially true when considering only variants that are further classified as pathogenic, likely pathogenic or VUS, using the ACMG classification. Further supporting the need to use the ACMG classification for stratification of the unknown *GBA1* variants is the heterogeneity that was observed before this stratification, and the lack of heterogeneity observed after the stratification.

As a group, carriers of ‘unknown’ *GBA1* variants may represent 1-1.5% of PD patients (Supplementary Data 2), which is not a negligible number. Currently, in most *GBA1*-targeting trials, these individuals are excluded due to their ‘unknown’ classification. Our results suggest that including carriers of *GBA1* variants with ‘unknown’ classification could be a feasible strategy to increase the number of trial participants, if they are also classified according to ACMG criteria as pathogenic, likely pathogenic or VUS. Based on the OR estimates, we would recommend currently considering these variants as equivalent to ‘risk’ variants if included in trials. That said, it is likely that some of these variants will be classified in the future as ‘severe’ or ‘mild’, if found in GD at a homozygous state or with a severe *GBA1* variant.

This study allows us to draw some conclusions about specific *GBA1* variants. The p.K(−27)R variant, is found within the 39 amino acids of the leader peptide of the glucocerebrosidase proenzyme. This variant is especially common in black populations, and the current study, as well as the lack of association of this variant with PD in a genome-wide association study of black individuals^11^ and the lack of GD patients homozygous to this variant, suggest that this is a benign variant that should not be included in *GBA1*-targeting trials. The p.E388K is often cited as a risk factor for PD.^12^ Indeed, in our meta-analysis, it was reported in seven studies with a total of 12,442 PD patients and 6,001 controls. There were a total of 12 (0.1%) PD patients and in 2 (0.03%) controls carrying this variant. However, in our single-variant meta-analysis, the p.E388K did not show a significant association, and additional studies are needed to determine if it is indeed a risk factor for PD. The p.A456P variant is often a part of recombinant alleles between the *GBA1* gene and its pseudogene, and since some studies may have failed to identify these recombinant alleles,^13^ we cannot comment about this specific variant. Other notable variants that were found only in patients are p.G(−1)R (4 patients), p.N392S (5) p.D453N (4), and p.I489V (6). The p.I(−20)V and p.R163Q variants are quite common and have similar frequencies in PD and controls, which argues for their exclusion from *GBA1*-targeting trials.

Our study has several limitations. It is based on studies with different methodologies for *GBA1* sequencing / genotyping which might affect some of the results. Since GWAS data was not available for these studies, we could not correct for ancestry principal components, yet our meta-analysis strategy partially mitigates this limitation, as most studies were done in ancestry-specific populations. Another limitation is that statistical power was severely restricted when the number of variants was too small. This made meaningful interpretation on specific variants difficult.

To conclude, we show that *GBA1* mutations classified as ‘unknown’ could be used for *GBA1*-targeting clinical trials with caution, currently considering them as ‘risk’ variants until they are properly classified.

## ACKNOWLEDGMENT

We gratefully acknowledge all individuals who contributed to the referenced clinical studies that form the foundation of this work. This work has been supported through grants from the Galen and Hilary Weston Foundation and the Michael J. Fox Foundation (MJFF). Additionally, the G-Can (*GBA1*-Canada) Initiative, an open-science collaborative initiative aimed at addressing *GBA1* mutation-based Parkinson’s disease, has made contributions to this research. G-Can is supported by The Hilary and Galen Weston Foundation, Silverstein Foundation, and J. Sebastian van Berkom and Ghislaine Saucier. S.C.P is supported by a fellowship from CIHR. Z.G.O is supported by the Fonds de recherche du Québec—Santé (FRQS) Chercheurs-boursiers award, and is a William Dawson Scholar.

## AUTHORS’ ROLES

S.C.P. was responsible for the conception, organization, and execution of the research project, as well as the design, acquisition, and analysis of the data, and wrote the first draft of the manuscript. Y.L. contributed to the data analysis and reviewed the manuscript. Z.G.O. contributed to the conception and organization of the research project, the design of the analysis, writing of the manuscript, and provided critical review and feedback.

