## Supplemental Figure for "*GBA1* variants with unknown classification are modest contributors to Parkinson’s disease susceptibility"

**Supplementary Figures 1 and 2**

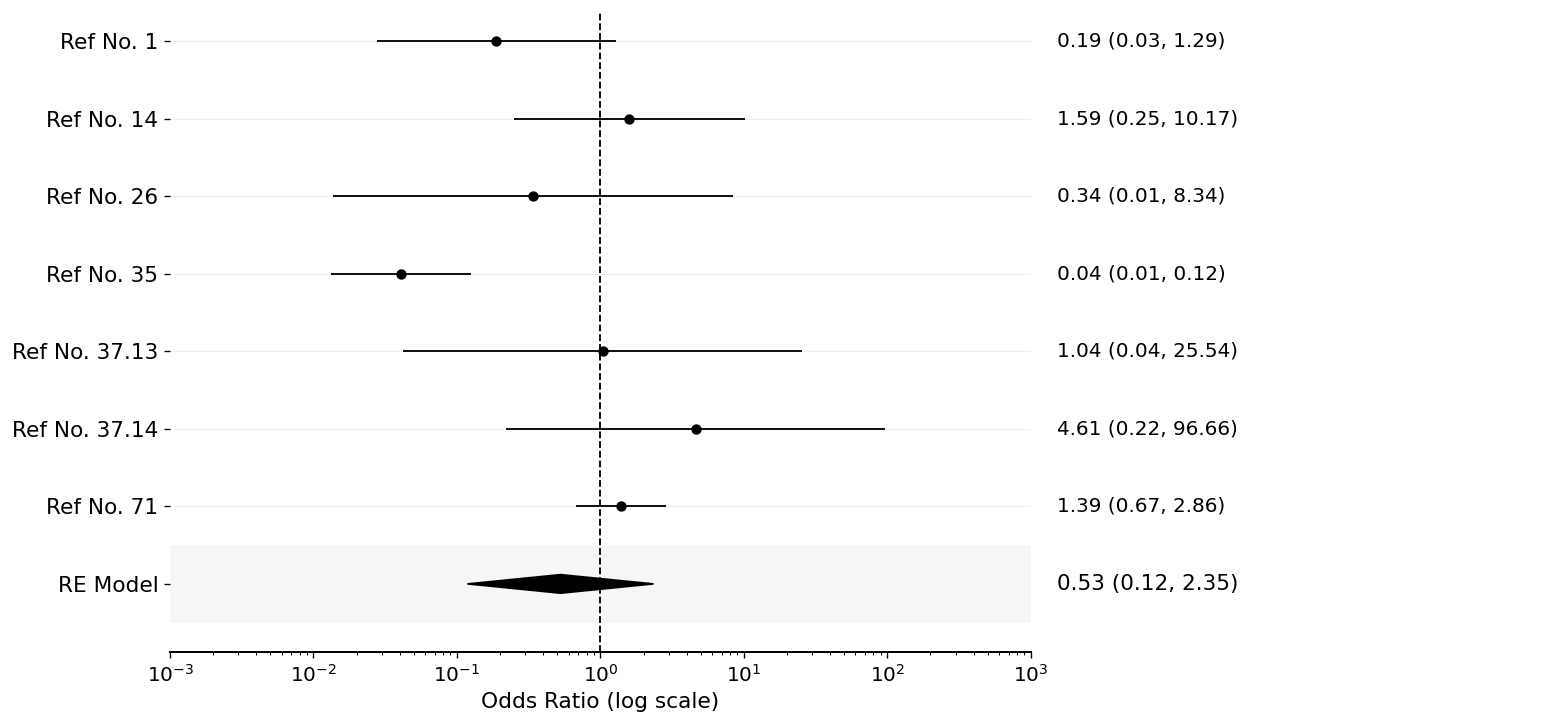

**Supplemental Figure 1.** Forest plot of studies with data on the *GBA1* p.K(-27)R variant. The pooled OR was 0.53 (95% CI: 0.12–2.35, p = 0.40), with moderate heterogeneity (Q=31.565, df=6, p(Q)=1.98 x 10^-5^, I²=81.0%).

RE = random effects; Ref No. = *GBA1*-PD browser reference number.

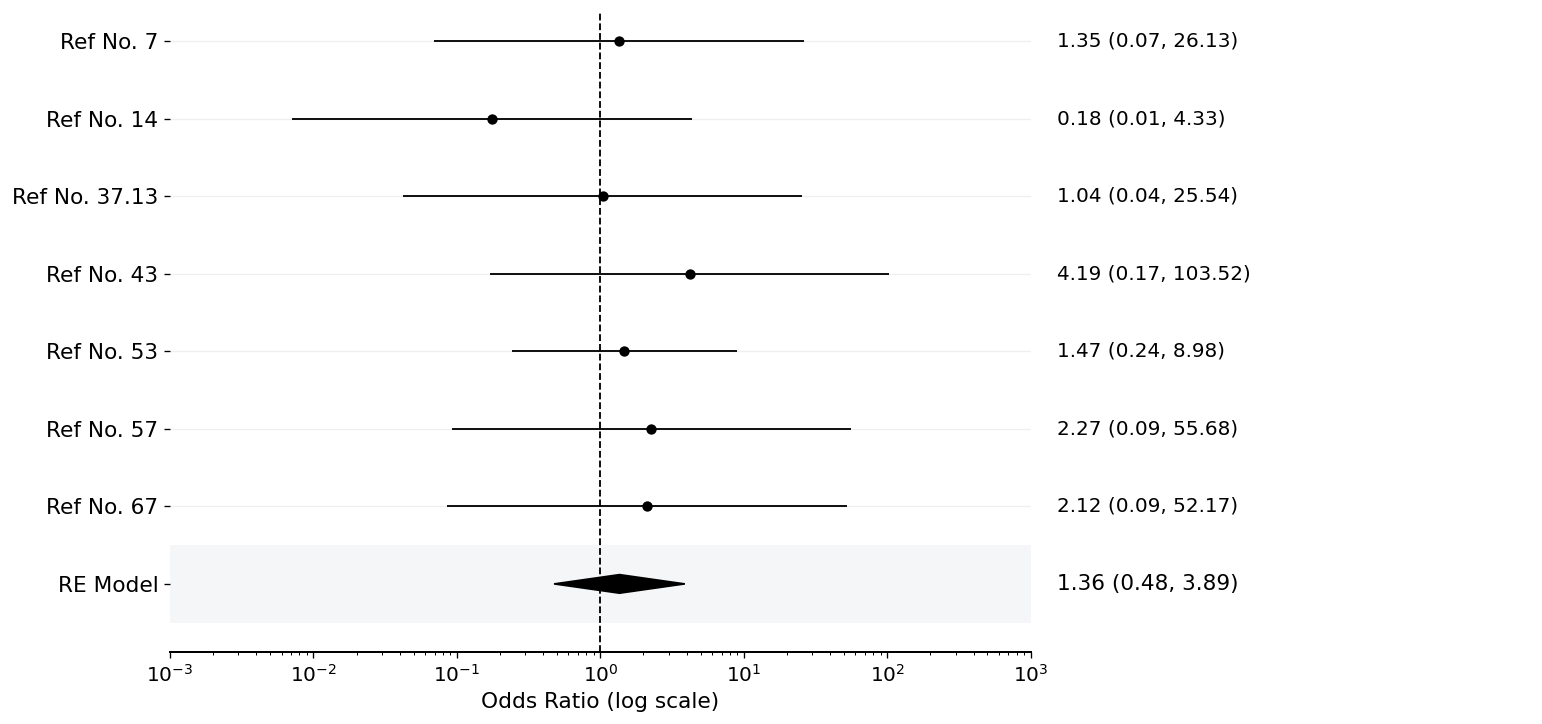

**Supplemental Figure 2.** Forest plot of studies with data on *GBA1* p.E388K variant. The pooled OR was 1.36 (95% CI: 0.48–3.89, p = 0.57), with no evidence of heterogeneity (Q=2.246, df=6, p(Q)=0.90, I²=0.0%)

RE = random effects; Ref No. = *GBA1*-PD browser reference number.
